# Machine-learning forecasting for Dengue epidemics - Comparing LSTM, Random Forest and Lasso regression

**DOI:** 10.1101/2020.01.23.20018556

**Authors:** Elisa Mussumeci, Flávio Codeço Coelho

**Affiliations:** Applied Mathematics School - Fundação Getulio Vargas - Rio de Janeiro - Brazil

**Author notes:** Corresponding author *Email address:* (Flávio Codeço Coelho).

**Keywords:** epidemiology, dengue, time series forecasting, LSTM, deep learning

## Abstract

Effective management of seasonal diseases such as dengue fever depends on timely deployment of control measures prior to the high transmission season. As the epidemic season fluctuates from year to year, the availability of accurate forecasts of incidence can be decisive in attaining control of such diseases. Obtaining such forecasts from classical time series models has proven a difficult task. Here we propose and compare machine learning models incorporating feature selection,such as LASSO and Random Forest regression with LSTM a deep recurrent neural network, to forecast weekly dengue incidence in 790 cities in Brazil. We use multivariate time-series as predictors and also utilize time series from similar cities to capture the spatial component of disease transmission. Among the compared models, the LSTM recurrent neural network model displayed the smallest predictive errors in predicting incidence of dengue out of sample, in cities of different sizes.

## 1. Introduction

Dengue is mosquito-borne viral disease which affects roughly 400 million people around the globe annually. Its transmitting vector, the Aedes mosquito (Mainly the aegypti and albopictus species), also transmits other serious viral diseases such as Zika, Chikungunya and Yellow fever, thus understanding the dynamics of Dengue can be beneficial the studies of other diseases transmitted by Aedes mosquitoes. Dengue is mostly restricted to tropical regions of the globe but its area of impact is expanding in response to global warming as well as to the accelerated growth of urban areas [1, 2]. Dengue is caused by 4 closely related serotypes of the dengue virus, which contributes to its endemicity since serotypes tend to alternate over time, and morbidity because secondary and subsequent dengue infections have a higher probability to evolve into a hemorrhagic fever.

Understanding and therefore being able to predict the incidence of mosquitoborne diseases is challenging due in part to the complex interplay between epidemiological and environmental determinants. This complex causal scenario manifests itself in the variability of incidence patterns in different geographical areas [2].The frequent lack of long-term historical records of disease incidence along with environmental variables affecting risk, further complicates statistical analysis.

In the case of dengue, the effects of climate on the vector’s population dynamics impose a marked seasonality which is then modulated by variations in the immunological structure of the population due to the co-circulation of multiple viral types, as well as other aspects of human demography: birth rates, immigration and short-term mobility.

Modeling dengue incidence patterns in Brazil presents an additional challenge characterized by the fact that the disease was reintroduced in the country in the 1980s having been absent for many decades. Therefore, besides the problem of short time series we are dealing with a disease which has yet to reach its endemic equilibrium. Any forecasting model assuming ergodicity will not perform well.

Instead of proposing parsimonious models built from previous knowledge of the determinants of dengue transmission, we adopted a machine learning approach, comparing models known for their ability to navigate their way into data intensive high dimensional problems by integrating variable selection into their fitting routine. Namely, we will compare the following machine-learning models: Long-Short-Term-Memory(*LSTM*), a recurrent deep neural network model designed to learn time-series, *Random Forest regression*(RF) and *LASSO regression*.

This approach has been tried before [3], but here we innovate by not only using a multivariate approach, but also in the way the predictor series are selected. We exploit the concept of epidemiological similarity to select the predictors for each city, and thus are able to compensate the short time span of our series with an ample set of similar series. Another distinguishing feature of this paper is that we fit and test the models in 790 Brazilian cities with a wide range of climate and demographic characteristics, exploring the robustness of each model.

## 2. Methodology

### 2.1. Data sources

The dataset used in the article was provided by the InfoDengue project. InfoDengue [4] is an integrated online dengue alert system. An unique source of carefully curated data for epidemiological studies, InfoDengue currently monitors 790 cities of 5 states in Brazil (Rio de Janeiro, Espirito Santo, Paraná, Minas gerais and Ceará), providing weekly analytical reports for Dengue, Zika and Chikungunya.

The data consist of weekly series(442 weeks in total) of dengue incidence, temperature (*°*C), relative humidity (%), atmospheric pressure(mmHg) and tweets about Dengue in each observed city. Meteorological data comes from local weather stations within each city. Twitter data was collected as described by de Almeida Marques-Toledo et al. [5]. It ranges from January 2010 until june of 2018. An excerpt of the data available for each city can be seen on figure 1.

**Figure 1:**
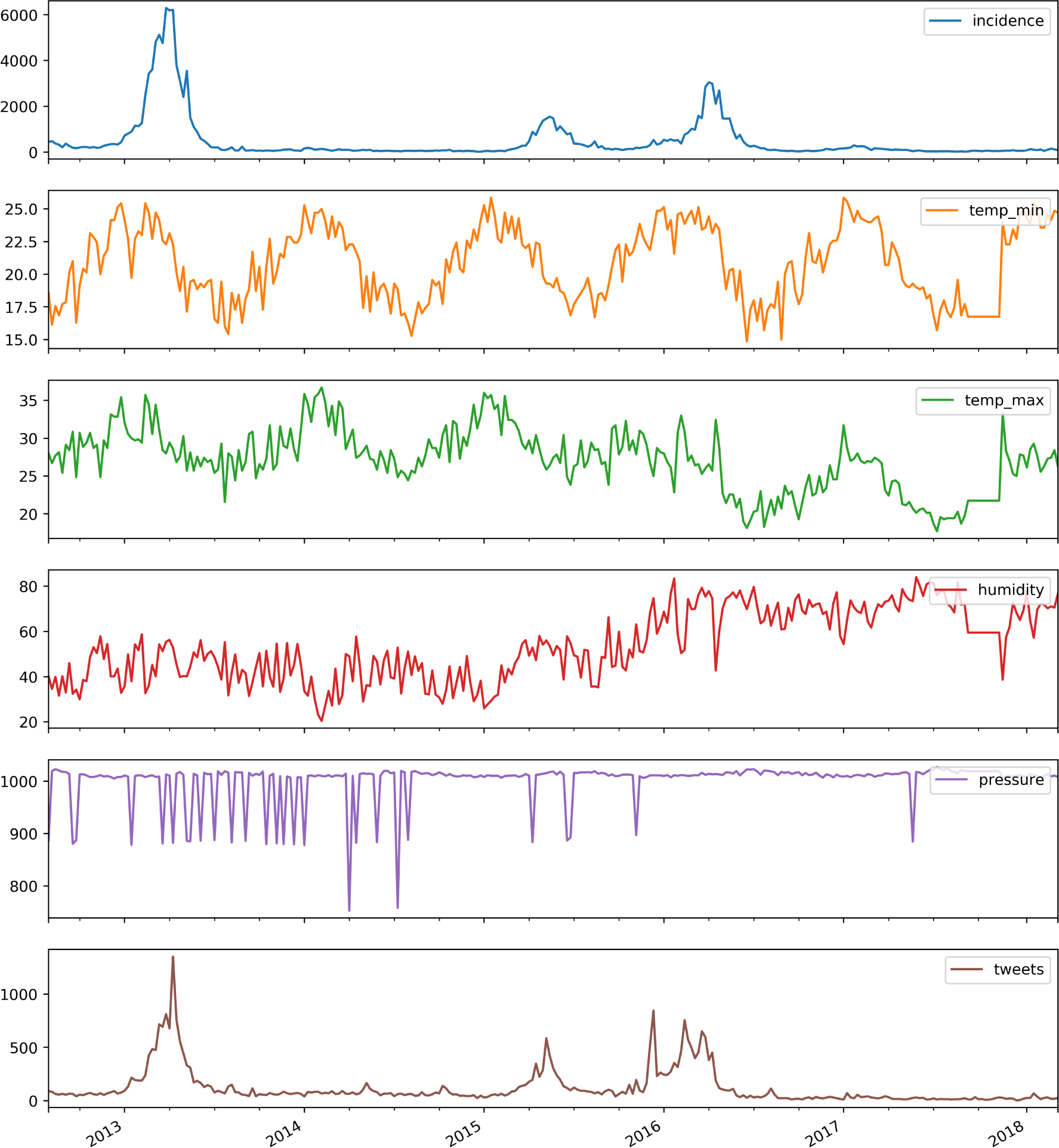
Time series available for every city in the dataset. The data shown here is from the city of Rio de Janeiro. incidence is shown as total reported cases per week; *temp_min* and *temp_max* are the minimum and maximum daily averaged over a week; *humidity* is show as average relative humidity at each week; *pressure* is the mean atmospheric pressure per week and finally, *tweets* is the number of tweets mentioning dengue per week.

### 2.2. Data pre-processing

It is very difficult to accurately forecast the weekly incidence in a city using only on its own historical data. This happens mainly because of the spatial component of disease transmission. A disease such as dengue relies on the movement of the virus among human sub-populations as a persistence mechanism, this flow is mostly maintained by human mobility patterns[6]. Thus, the flow of individuals between cities can also be an important predictive factor for incidence.

To tap into the spatial component of dengue dynamics [7, 8], it makes sense to uses series from neighboring cities to inform on the local incidence. Additionally, other cities not necessarily in the vicinity but which display similar historical series of incidence, can also be included as predictors. Besides, the flow of people between cities is not restricted to contiguous neighbors.

In order to define the set of cities with relevant predictive information for each city, we clustered all cities within a state based on the correlation distances(eq. 1) between the incidence time series for each pair (*u, v*) of cities.

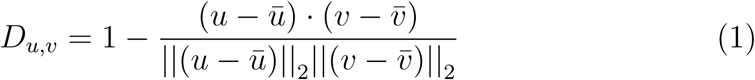

above, 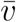 and 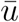 are the mean of the series for city *v* and *u*, respectively, and the product in the numerator is the dot product.

The clusters were calculated hierarchically, where the distance *d* between each pair of clusters (*c*_1_, *c*_2_) was given by the “Farthest Point Algorithm”, *d*(*c*_1_, *c*_2_) = *max*(*dist*(*c*_1_[*i*], *c*_2_[*j*])), for all points *i* in cluster *c*_1_ and *j* in cluster *c*_2_. The threshold is set to 0.6 *× max*(*Z*), where *Z* is the vector of the pairwise Spearman correlation distances between the cities.

In order to train the model for each city *i*, a feature matrix *X*^*i*^, composed of incidences, minimum temperatures, maximum temperatures, relative humidities and atmospheric pressure time-series, was assembled from the set of cities in its cluster. Figure 2 shows an example of this clustered feature matrix.

**Figure 2:**
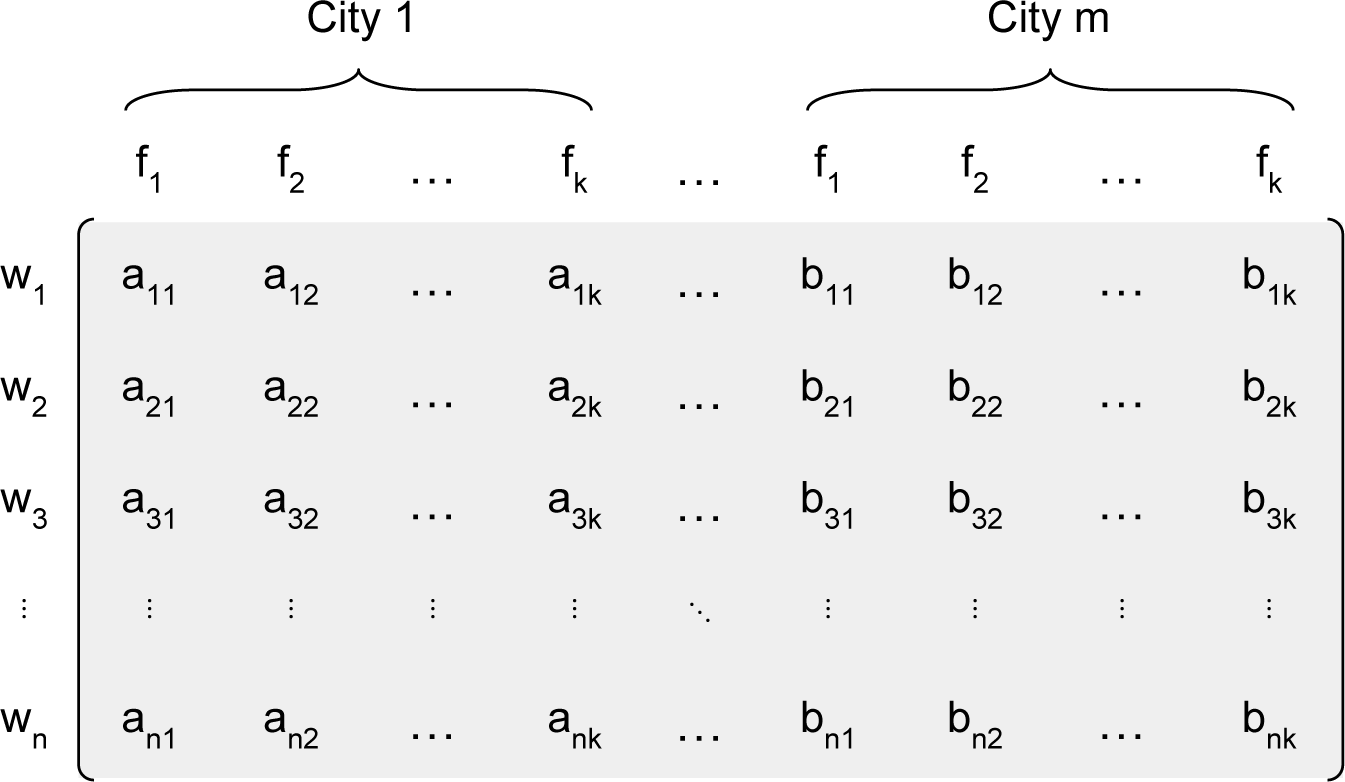
Clustered features data with *n* as the total number of weekly observations, *m* as the total number of cities belonging to a cluster and *k* the total number of series(features) per city.

The clustering was done within each state, i.e., cities can only be clustered with others from the same state.

To serve as a baseline for the effectiveness of using a cluster of cities as predictors, for each type of model described below, we also trained them without variables from the cluster.

### 2.3. Forecasting

The general problem we try to solve here is that of predicting future incidence in city *i* using past states of itself and the set of predictors composing matrix *X*^*i*^. We train tree different classes of models (detailed below) to forecast the absolute dengue incidence time-series, i.e, the number of cases reported each week, as shown in fig. 1. In all models we adopted a forecast window of 4 weeks, meaning that from any moment in time the models will produce forecasts for the number of weekly dengue cases in the following 4 weeks, based only on historical data up to that point.

#### 2.3.1. Random Forest regression

We used a Random Forest regression model to predict a single point in the future based on historical data. In order to turn a time series prediction problem into a random forest regression model, we transformed the series regressors into a vector *T* containing the 𝒟 most recent observations from each series in the feature matrix *X*.

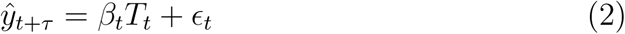

here *T*_*t*_ is defined as *T*_*t*_ = [*X*_*t*_, *X*_*t−*1_, …, *X*_*t−D*_]^*T*^, where *X*_*t−d*_ is the vector with the values of all *mk* (*k* series for each of the *m* city in the cluster) predictor series at time *t − d*, where *d* = 0, …, *D. β*_*t*_ is a vector 1 *× mk* which contains the weights for each value of *T*_*t*_. The model predicts the incidence at a particular week in the future *y*_*t*+*τ*_, thus since we wanted to predict 4 weeks into the future, 4 separate models were fitted to data for each *τ* varying from 1 to 4.

Figure 3 illustrates the data transformations required by the Random Forest Regression model.

**Figure 3:**
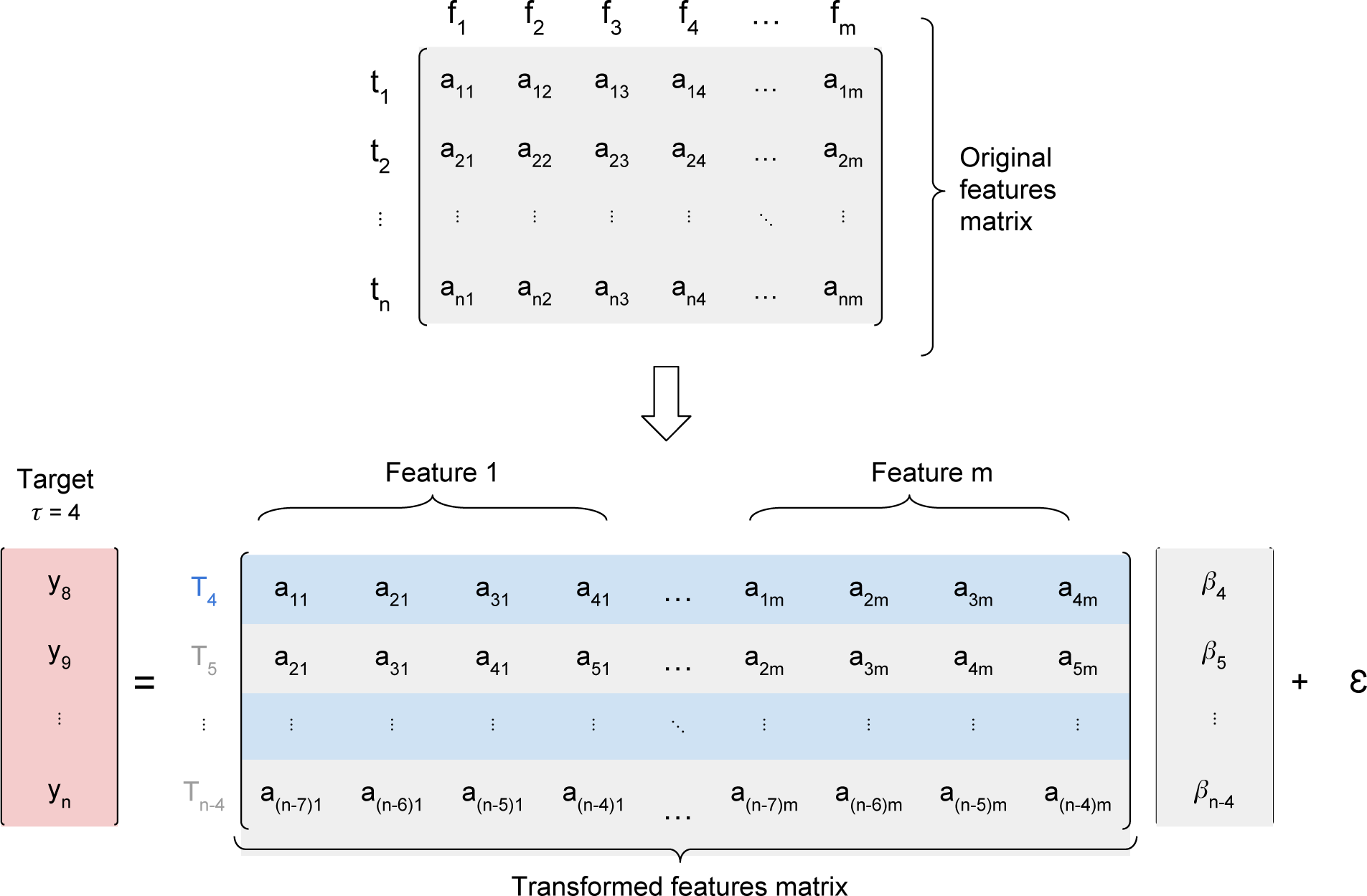
Feature matrix transformation for Random Forest input. The first matrix is the original data, where *a*_*ij*_ is the feature *i* at time *j, j* = [1, ‥, *n*], and *y*_*i*_ is the incidence of dengue cases at time *i, i* = [1, …, *n*]. In this example, *D* = 4 and *τ* = 4, so our first vector is *T*_4_, which contains all m predictors series at times (*t*_1_, *t*_2_, *t*_3_, *t*_4_), and our target is *y*_*t*+*τ*_ = *y*_*D*+*τ*_ = *y*_8_. The last input must contain the last target, therefore *y*_*t*+*τ*_ = *y*_*n*_ and *T*_*t*_ = *T*_*n−*4_.

##### Long Short Term Memory (LSTM)

A LSTM model is a recurrent deep neural network model developed to handle predictions of timeseries. We used a LSTM model with topology given in table 1. The model was trained for 300 epochs using a mean-log squared-error (MLSE) loss function (eq. 3) and a Nesterov Adam optimizer[9]. Again, as in the RF model, the 4 most recent weeks were used as predictors, and a forecasting window of 4 weeks was chosen. The input tensor fed into the network is described in figure 4.

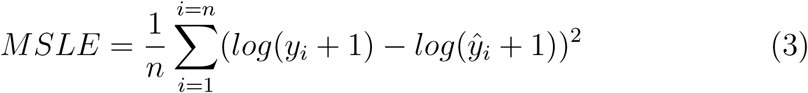

**Table 1:** Topology of LSTM neural network. There are three LSTM layers with 0.2 dropout between them and a dense layers which gives the incidences in the following 4 weeks.

| Layer | Output | Activation | Dropout |
| --- | --- | --- | --- |
| LSTM | (1,4,4) | tanh | 0.2 |
| LSTM | (1,4,4) | tanh | 0.2 |
| LSTM | (1,4) | tanh | 0.2 |
| Dense | (1,4) | relu | - |

**Figure 4:**
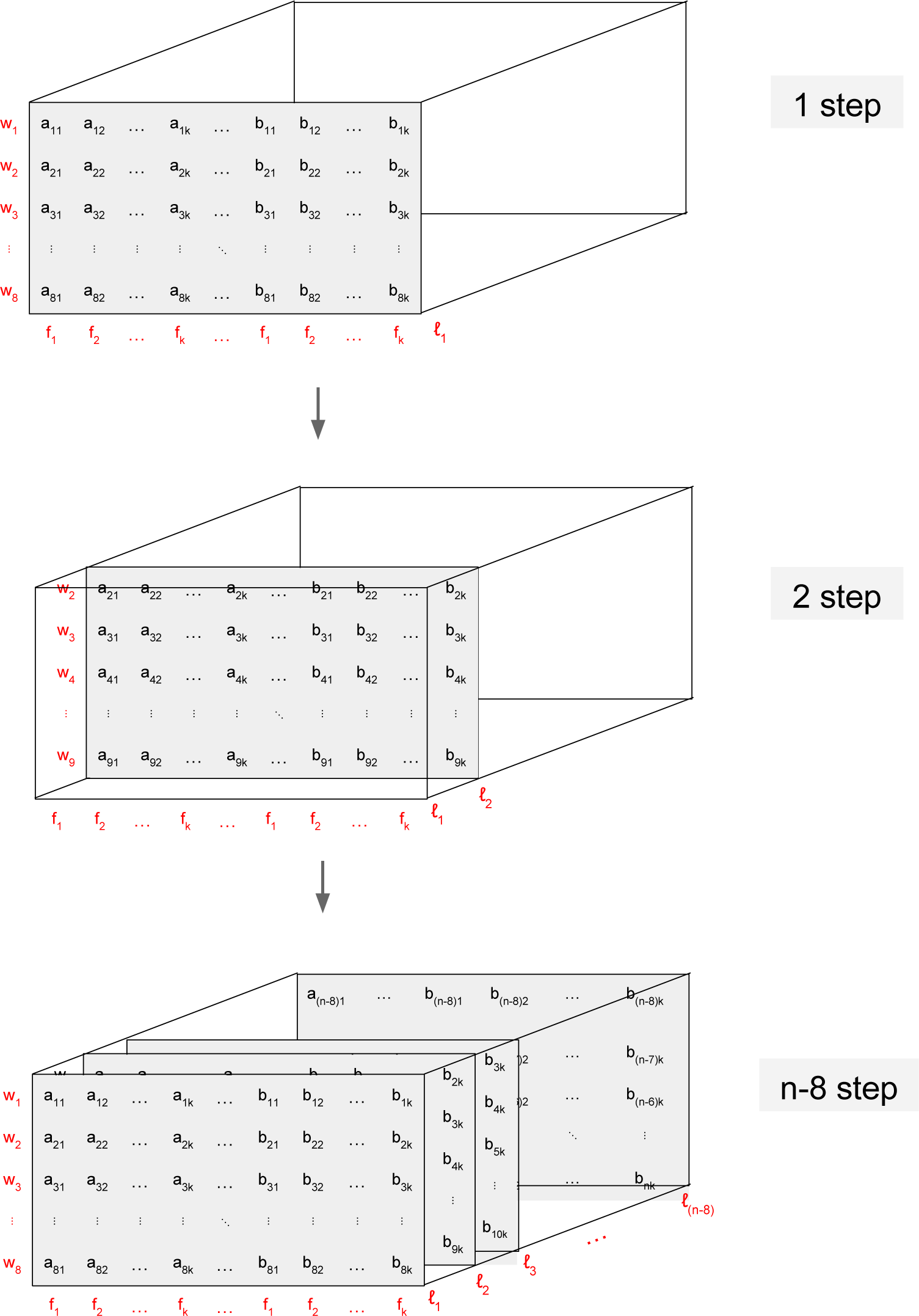
Structure of the input tensor used to train the LSTM model.

The input tensor LSTM is 3-dimensional(2). The first dimension of the matrix is the number of features; the second dimension is the length of series (total number of weeks - (look-back + forecasting window)); the third dimension is the length of the moving window fed to the model (look-back + forecasting window).

The figure 4 illustrates the structure of this tensor when forecasting window equal to 4 and look-back window *D* = 4.

##### LASSO regression

The third model evaluated was a LASSO least angle regression[10]. The same feature matrix used for the Random Forest regression applies here. Similarly, 4 separate models were fit, one for each value of *τ*.

##### Validation

Validation of the forecasts must take into account that the data points which are the targets of the forecast, are subject to observational errors and natural stochasticity. So we defined the ground truth for validating the model the percentile of the historical observations for every week and every city. Let *H*_*i,w*_ be the empirical distribution of incidence on city *i* and week *w* and *P*_*i,w*_, represent the percentile of *H*_*i,w*_ to which the observed incidence at week *w* at city *i* corresponds to. Similarly, Let 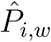 represent the percentile of the model’s prediction. A good agreement between model and data would lead to their percentiles being very similar for most points. Deviations from equality would indicate under- or over-estimation issues. By mapping the data and predictions to a percentile, allow us to pool all predictions from all weeks and cities into a comparable scale. Using this scale we can validate the performance of the models regardless the original scale of the data, and year-to-year differences in magnitude.

## 3. Results

### Cluster analysis

Figure 5 shows the incidence series for one cluster for the entire historical period available. We can see that cities clustered together display similar incidence patterns. Number of clusters per state and average sizes are given on table 2.

**Table 2:** Cluster characteristics for each state.

|  | Number of cities | Number of Clusters | Cluster mean size |
| --- | --- | --- | --- |
| **Rio de Janeiro** | 92 | 14 | 6 |
| **Paraná** | 399 | 54 | 6 |
| **Ceará** | 184 | 49 | 3 |

**Figure 5:**
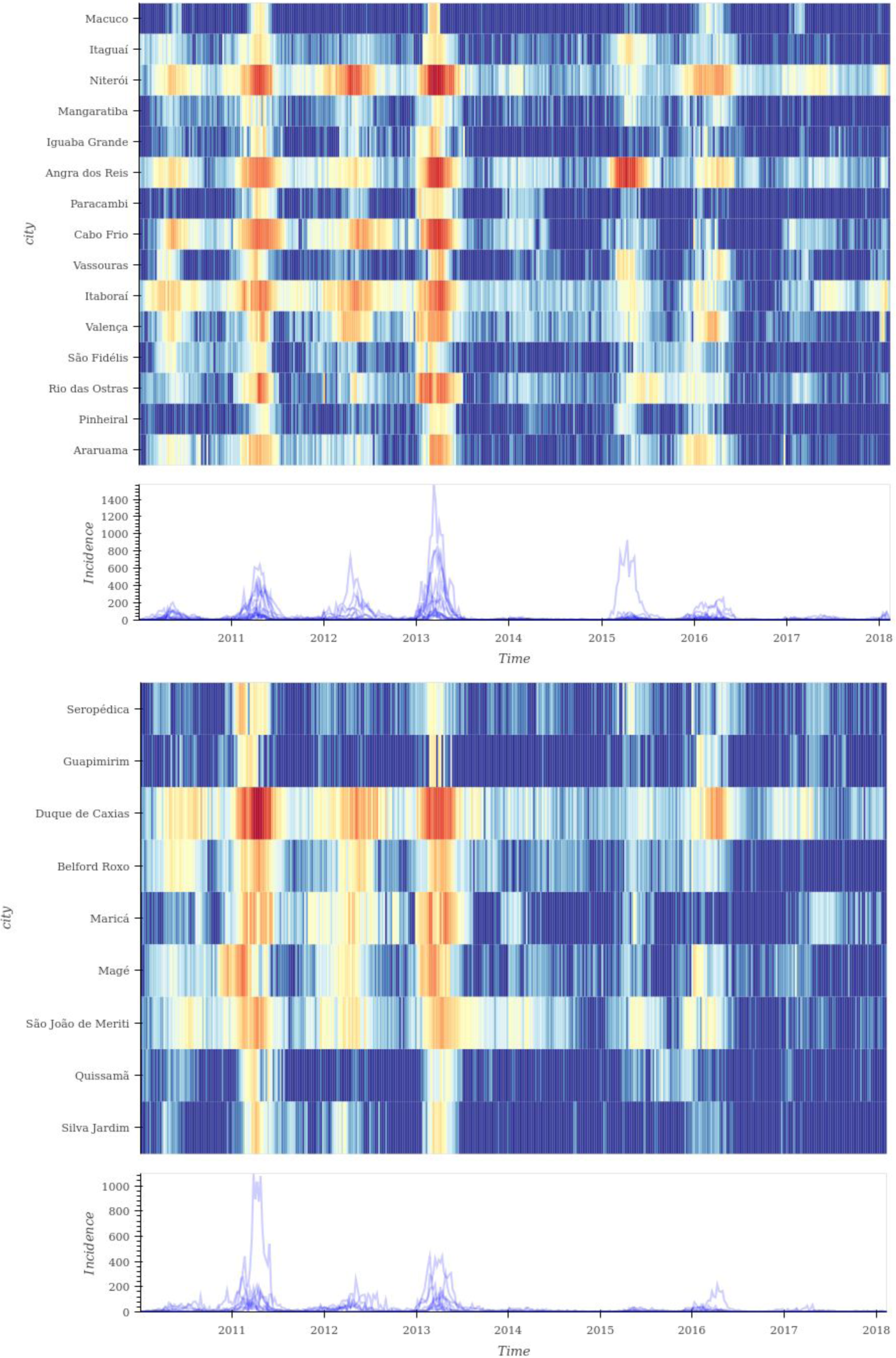
Dengue incidence for two clusters from Rio de Janeiro state. The colors represent the incidence of dengue through time, with red being the highest incidence dark blue the lowest.

On figure 6, we can see the clustering in the Rio de Janeiro state.

**Figure 6:**
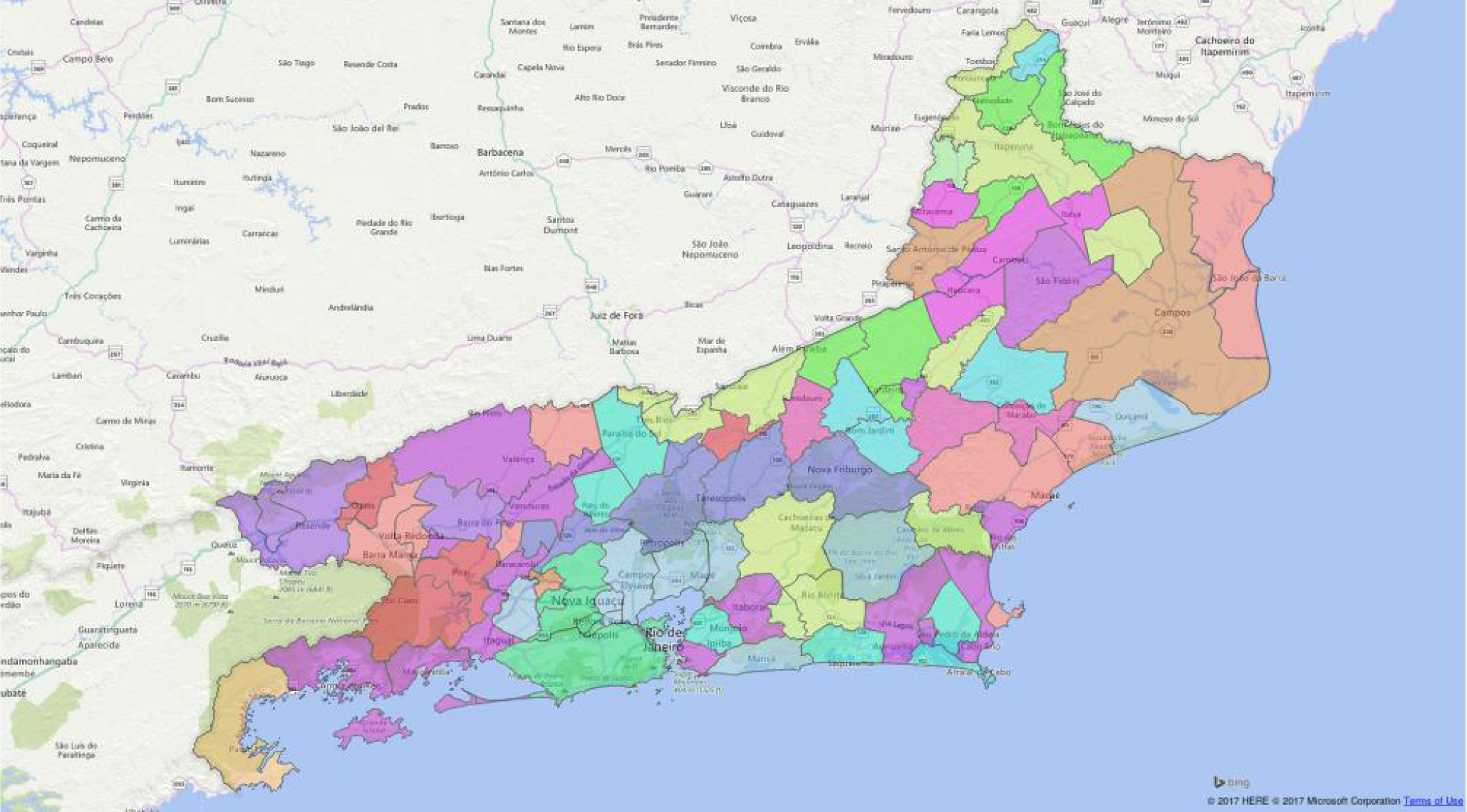
Map representing clusters of cities in Rio de Janeiro state, through color coding. Cities with the same color belong to the same cluster. Notice how clusters are not contiguous geographically.

### Forecasting

We measured the performance of the forecasting models by the magnitude of the prediction error for week *t* + 4 where *t* is the last week observed. The prediction errors were calculated as mean squared error (MSE), and mean squared log error(MSLE).

Figures 7 and 8 show the performance of the forecast both *in-sample* and *out-of-sample*. On figure 7, we see the forecast for 2 cities and the three models: For *Rio de Janeiro*, all 3 models seem to do an equally good job at predicting out of sample while for *Campos dos Goytacazes*, they all seem to struggle. Over the entire set of cities, LSTM and Random Forest performed better as shown on figure 9. Table 3, shows the mean errors for all models.

**Table 3:** Mean prediction errors in quantile scale 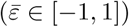 and standard deviations for all models in-sample(in) and out-of-sample(out).

| Model | RJ |  | CE |  | PR |  |
| --- | --- | --- | --- | --- | --- | --- |
| data | in | out | in | out | in | out |
| **Lasso** | $0.4 \pm 0.45$ | $0.53 \pm 0.38$ | $0.4 \pm 0.41$ | $0.33 \pm 0.50$ | $0.45 \pm 0.44$ | $0.47 \pm 0.42$ |
| **RF** | $0.17 \pm 0.30$ | $0.33 \pm 0.38$ | $0.13 \pm 0.27$ | $0.3 \pm 0.47$ | $0.18 \pm 0.3$ | $0.39 \pm 0.42$ |
| **LSTM** | $-0.06 \pm 0.41$ | $0.04 \pm 0.35$ | $0 \pm 0.34$ | $-0.07 \pm 0.45$ | $-0.08 \pm 0.36$ | $-0.05 \pm 0.35$ |

**Figure 7:**
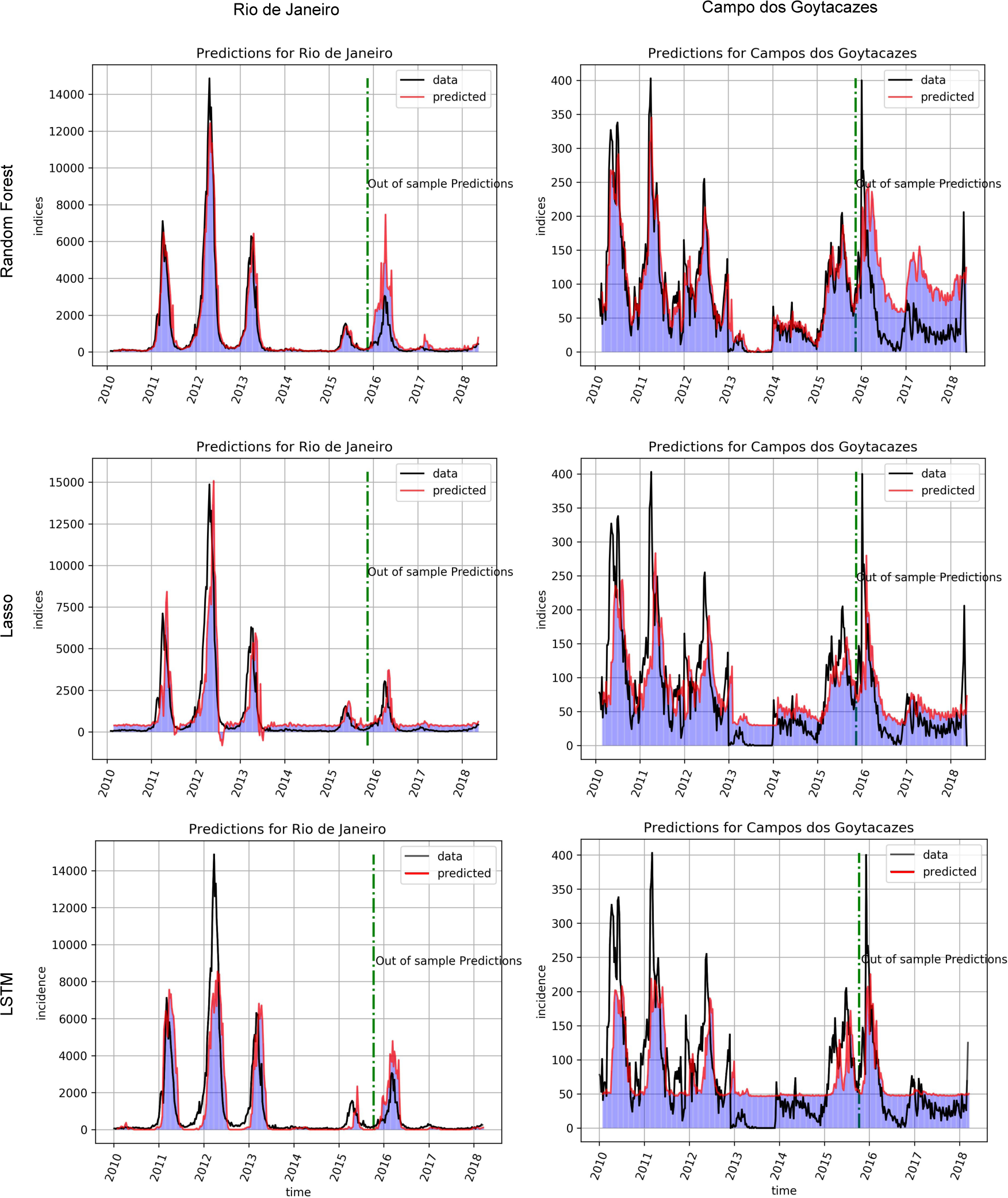
In-sample and out-of-sample forecast prediction for *Rio de Janeiro* and *Campos dos Goytacazes*. The black line is the observed data and the redline represents predictions 4 weeks after the last observed point.

**Figure 8:**
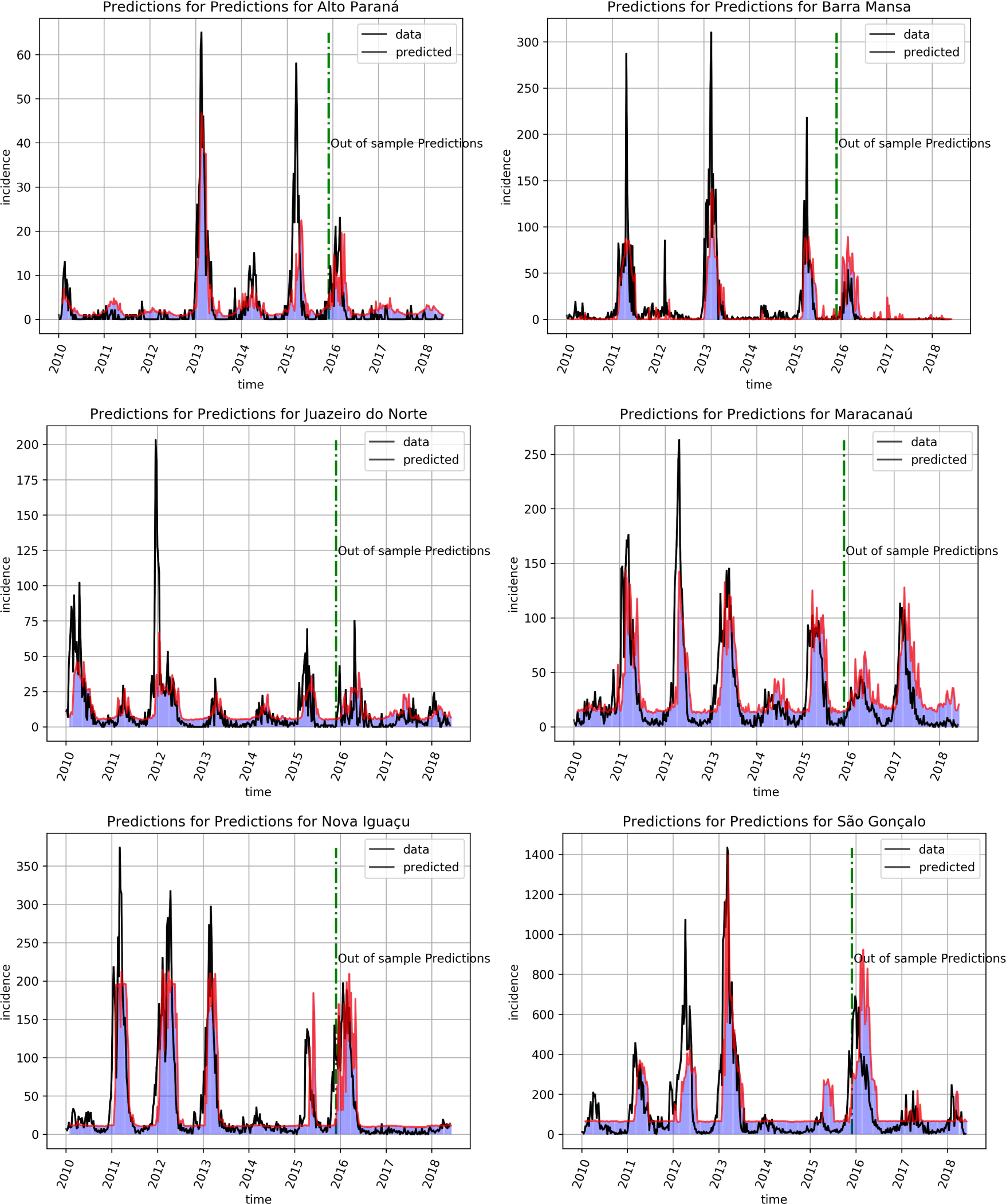
Results for six other cities with varying sizes and from different regions of the country: *Alto Paraná* from the south, *Barra Mansa, São Gonçalo* and *Nova Iguaçú* from the southeast and the others from the Northeast.

**Figure 9:**
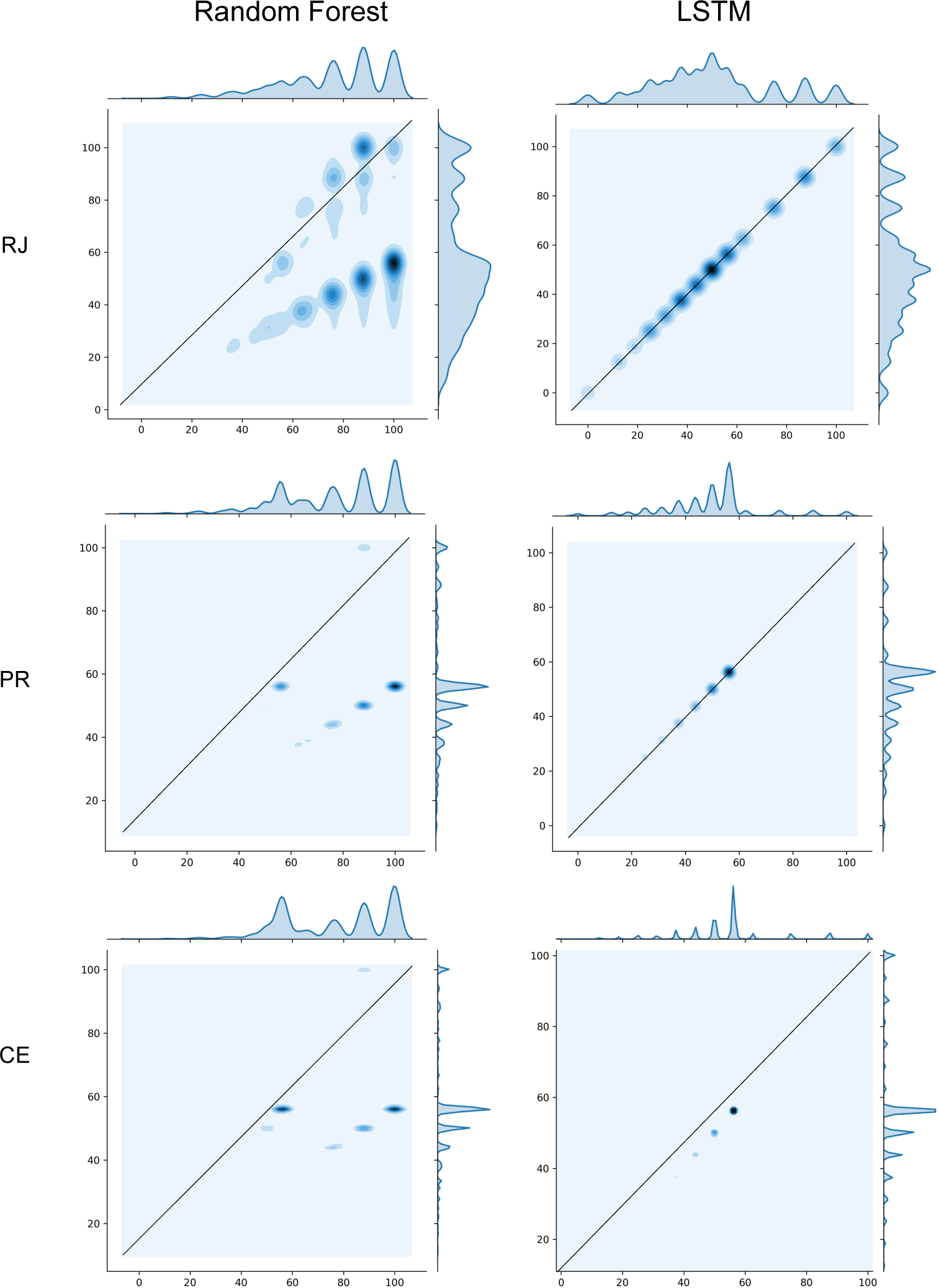
In the panels above, we show observed vs predicted distributions for all out of sample forecasts generated for all cities grouped by state. The *x*-axis encodes the percentiles of the predicted points distribution, while the *y*-axis are the percentiles of the distribution of observed values. Each point (*x, y*) giving rise to the heatmaps, represents one prediction and one expected observation1. 9If the predicted value and the observed value of a week *i* is near the line *x* = *y*, the predicted value is consistent with the historically expected for that city in that week. We plotted the density of the points, so the dark blue areas are where the most values belongs to.

Figure 8 shows forecasts for 6 other cities generated by the LSTM model.

## 4. Discussion

From the results obtained for the three models evaluated in this paper, we realized that machine learning models show a great potential to be used in the forecasting of dengue incidence time series. Their ability to deal with the non-linearities as well as with the non-stationary and heavy-tailed distributions of weekly incidences is a major advantage compared to classical auto-regressive models[11].

The two regression-based models, RF and LASSO, have the advantage of being computationally less costly than LSTM, which can be very important for the application described here which is to provide forecasting for hundreds of cities on a weekly basis, as is the case with the Infodengue project. RF and LASSO take on the order of seconds (on a 32-core CPU) to train on our full dataset, the LSTM model, on the other hand takes close to ten minutes to train on the same dataset, but on a Nvidia Tesla K40 GPU with 2880 cores. When we have to train hundreds of models, the computational cost of LSTM can get very high. However, even though is desirable to update the models weekly, our out-of-sample results have shown that the models remain accurate for over two years without retraining.

The accuracy winner in this comparison was the LSTM model which was able to better approximate the empirical distribution of dengue’s weekly incidence as can be seen in the marginal distributions of figure 9. LSTM was also the least biased of the three models slightly overestimating the very low incidences of interepidemic periods while underestimating high epidemic peaks (see figures 8 and 9).

There are many ways in which the performance of the models compared here can still be improved. In the present study we maintained the design of the models as standard as possible as a first approach to the problem. In the clustering of the cities it can be useful to consider lagged correlations between series as the causal dependencies between cities are not likely to be instantaneous. Differentiation and log transforming of the series is also likely to be interesting as it would stabilize the distribution helping the models to deal with extreme values.

In the deep learning field, many variations to the standard LSTM model have been proposed[12, 13] which could be tested with this dataset. Recent methodologies [14] to assess the uncertainty of deep learning model can be applied to this model.

## 5. Conclusion

The results presented here add to the still scarce literature of the applicability of machine learning models to epidemiological forecasting[15, 3, 16]. We have demonstrated that the Deep learning models such as the LSTM can be used with good performance in large scale predictive problems. The Infodengue project is going to integrate the LSTM model to provide forecasts of dengue incidence in Brazilian cities and we hope to see a wider evaluation and adoption of machine-learning models in this setting.

## Data Availability

Data used in this paper is available through the Infodengue Project API in the link given below.

https://info.dengue.mat.br/services/api

## References

[1] R. T. Watson, J. Patz, D. J. Gubler, E. A. Parson, J. H. Vincent, Environmental health implications of global climate change, Journal of Environmental Monitoring 7 (2005) 834–843.

[2] S. Bhatt, P. W. Gething, O. J. Brady, J. P. Messina, A. W. Farlow, C. L. Moyes, J. M. Drake, J. S. Brownstein, A. G. Hoen, O. Sankoh, et al., The global distribution and burden of dengue, Nature 496 (2013) 504.

[3] Y. Shi, X. Liu, S.-Y. Kok, J. Rajarethinam, S. Liang, G. Yap, C.-S. Chong, K.-S. Lee, S. S. Tan, C. K. Y. Chin, et al., Three-month real-time dengue forecast models: An early warning system for outbreak alerts and policy decision support in singapore, Environmental Health Perspectives 124 (2016) 1369.

[4] C. Codeco, O. Cruz, T. I. Riback, C. M. Degener, M. F. Gomes, D. Villela, L. Bastos, S. Camargo, V. Saraceni, M. C. F. Lemos, F. C. Coelho, Infodengue: a nowcasting system for the surveillance of dengue fever transmission, bioRxiv (2016).

[5] C. de Almeida Marques-Toledo, C. M. Degener, L. Vinhal, G. Coelho, W. Meira, C. T. Codeço, M. M. Teixeira, Dengue prediction by the web: Tweets are a useful tool for estimating and forecasting dengue at country and city level, PLoS neglected tropical diseases 11 (2017) e0005729.

[6] B. Adams, D. D. Kapan, Man bites mosquito: understanding the contribution of human movement to vector-borne disease dynamics, PloS one 4 (2009) e6763.

[7] S. T. Stoddard, B. M. Forshey, A. C. Morrison, V. A. Paz-Soldan, G. M. Vazquez-Prokopec, H. Astete, R. C. Reiner, S. Vilcarromero, J. P. Elder, E. S. Halsey, et al., House-to-house human movement drives dengue virus transmission, Proceedings of the National Academy of Sciences 110 (2013) 994–999.

[8] L. Eisen, S. Lozano-Fuentes, Use of mapping and spatial and space-time modeling approaches in operational control of aedes aegypti and dengue, PLoS Negl Trop Dis 3 (2009) e411.

[9] I. Sutskever, J. Martens, G. Dahl, G. Hinton, On the importance of initialization and momentum in deep learning, in: International conference on machine learning, pp. 1139–1147.

[10] B. Efron, T. Hastie, I. Johnstone, R. Tibshirani, et al., Least angle regression, The Annals of statistics 32 (2004) 407–499.

[11] C. Chatfield, Time-series forecasting, Chapman and Hall/CRC, 2000.

[12] N. Laptev, J. Yosinski, L. E. Li, S. Smyl, Time-series extreme event forecasting with neural networks at uber, in: International Conference on Machine Learning, 34, pp. 1–5.

[13] J. Cao, Z. Li, J. Li, Financial time series forecasting model based on ceemdan and lstm, Physica A: Statistical Mechanics and its Applications 519 (2019) 127–139.

[14] Y. Gal, Z. Ghahramani, Dropout as a bayesian approximation: Representing model uncertainty in deep learning, in: international conference on machine learning, pp. 1050–1059.

[15] Y. L. Hii, H. Zhu, N. Ng, L. C. Ng, J. Rocklöv, Forecast of dengue incidence using temperature and rainfall, PLoS neglected tropical diseases 6 (2012) e1908.

[16] X. Zhang, T. Zhang, A. A. Young, X. Li, Applications and comparisons of four time series models in epidemiological surveillance data, PLoS One 9 (2014) e88075.

